# Using Trends and Outliers in Managing Delayed Transfusions

**DOI:** 10.1101/2022.10.03.22279993

**Authors:** Burak Bahar, Eric A. Gehrie, Yunchuan D. Mo, Cyril Jacquot, Meghan Delaney

**Author notes:** Co-first authors (Burak Bahar, Eric A. Gehrie). Corresponding author: Burak Bahar, MD, Children’s National Hospital, 111 Michigan Ave NW, Department of Laboratory Medicine, Washington, DC, USA.

## Abstract

Delayed transfusions may result in patient morbidity and mortality, but no standards for timely transfusion have been developed. Information technology tools could be implemented to identify gaps in provision of blood and to recognize areas of improvement. Herein, we propose that the time to initiate a blood transfusion after an informative laboratory test could feasibly be used by the transfusion medicine service as a metric to monitor for transfusion delays. Trends and outlier events could be further investigated and used to make decisions and implement protocols to improve patient care.

## Introduction

An emerging area of focus within transfusion medicine is the possibility that efforts to curtail unnecessary blood transfusions could unintentionally deprive patients of needed blood transfusions (1). However, the ability to measure or detect transfusions that should have occurred – but did not – is frustrated by the fact that transfusion medicine services generally begin their review at the time of order receipt. In addition, blood shortages could deprive a patient of a needed transfusion even if the clinical indication is recognized by the clinical service (1). The Serious Hazard of Transfusion (SHOT) program defines delayed transfusion as “where a transfusion of a blood component was clinically indicated but was not undertaken or non-availability of blood components led to a significant delay (*e*.*g*., that caused patient harm, resulted in admission to ward or return on another occasion for transfusion)” and accepts reports of such events (2). Since 2011, there has been is an increasing trend in the number of annual delayed transfusion reports (3). Per the SHOT 2021 Annual Report, there were 179 delayed transfusion reports and nine deaths in the UK in 2021 (3). SHOT recommended actions to prevent transfusion delays include reviewing, updating and implementing policies, implementing training programs and processes to audit and investigate all transfusion delays, and using appropriate investigation tools (4). We propose that the average (or median) time to initiate a blood transfusion after an informative laboratory test (*e*.*g*., haemoglobin concentration or platelet count below local threshold) could be used by the transfusion medicine service as an investigation tool to assess transfusion delays. This metric could also be used to evaluate trends and identify outlier events. This knowledge could help to implement approaches to improve stewardship of the blood supply and minimize the impact of blood shortages.

## Methods

From January 1^st^, 2020, to September 1^st^, 2022, all patients’ complete blood cell (CBC) result and red blood cell (RBC) and platelet transfusion data were extracted, transformed, and loaded into our previously described laboratory data science platform (https://github.com/bbahar/umay) (5) with additional custom R and SQL scripts (https://github.com/bbahar/delayed-transfusions). The number of RBC and platelet transfusions per week were calculated. Time from the release of laboratory results to the initiation of transfusions were calculated and weekly medians were used for trend analyses. Variables of interest for RBC transfusions were haemoglobin level (<7g/dL and ≥7g/dL), RBC minor antibody presence and Rh status of the recipient, and for platelet transfusion was the platelet count (≤10k/μL, >10 to ≤50k/μL and >50 to ≤150k/μL).

Outliers in the time series data were detected utilizing the ‘anomalize’ R package (version 0.2.2) on R (version 4.0.5). To accommodate for seasonal and trend changes and to find the outliers, data were decomposed using locally estimated scatterplot smoothing (LOESS) and turned into remainder values (*R*) which were compared to their remainder limits (*Rlim*) obtained using generalized extreme studentized deviate (GESD) test. *R* and *Rlim* were calculated using default alpha (width of the normal range), frequency and trend window settings (0.05, 12 to 13 weeks and 30 to 45 weeks, respectively). Maximum number of outliers permitted were 2 percent of the data. Results are expressed in mean (standard deviation [SD]) or median (minimum [min], maximum [max]), as appropriate. The study was conducted under Children’s National Hospital Institutional Review Board approved study protocol (Pro00010083-Mod00000781 ‘effectiveness of blood product transfusion and associated adverse events in neonatal & paediatric patients’). A two-tailed Student’s *t*-test was performed to assess for significance for continuous variables.

## Results

Over the two years and eight months period, 1,587 patients received 10,946 RBC transfusions and 730 patients received 7,704 platelet transfusions within 24 hours of a haemoglobin or a platelet count result, and patients were aged 11.1 years (SD: 7.03) and 10.8 years (SD: 7.03), respectively. Average volume of RBC transfusion was 251.7 mL (SD: 95.8), average platelet transfusion volume was 148.9 mL (SD: 79.2), and time to initiate a transfusion was 11.1 hours (SD: 6.82) and 11.1 hours (SD: 7.14), respectively. Mean time to initiate a transfusion was shorter for patients with haemoglobin <7 g/dL (8.67 hours [SD: 5.46]) and platelet count ≤10k/μL (7.52 hours [SD: 6.3]) than haemoglobin ≥7 g/dL (11.83 hours [SD: 6.99]) and platelet count >10k/μL (11.76 hours [SD: 7.09]) (both *P*: <0.001). On average there were 79 (SD: 25) RBC and 55 (SD: 20) platelet transfusions per week.

Dates of outliers for the median time to initiate an RBC or a platelet transfusion after a CBC result were reported in Table 1 and were visualized in Figure 1. Overall, aggregated data assessment demonstrated that there was one outlier week (2020-10-12 to 2020-10-18, *R*: 7.48 [*Rlim: -*6.53 — 6.44]) for all RBC transfusions and none for all platelet transfusions.

**Table 1.** Dates of outliers for weekly median time to initiate a red blood cell or a platelet transfusion following a haemoglobin, or a platelet count result and based on the haemoglobin level, red blood cell minor antibody presence, Rh status, and platelet count of the patient.

| Variable |  | Dates of outliers | *R* | *Rlim* |
| --- | --- | --- | --- | --- |
| All RBC transfusions |  | 2020-10-12 to 2020-10-18 | 7.48 | -6.53 — 6.44 |
| RBC transfusions based on haemoglobin level |  |  |  |  |
|  | < 7g/dL | 2020-07-20 to 2020-07-26 | 6.0 | -5.46 — 5.61 |
|  | ≥ 7g/dL | None |  |  |
| RBC transfusions and antibody presence |  |  |  |  |
|  | Antibody positive | None |  |  |
|  | Antibody negative | 2021-08-09 to 2021-08-15 | 7.03 | -6.47 — 6.13 |
|  |  | 2022-07-11 to 2022-07-17 | -7.31 | -6.47 — 6.13 |
| RBC transfusion and Rh status of recipient |  |  |  |  |
|  | Rh positive | 2020-10-12 to 2021-08-15 | 7.84 | -7.38 — 7.07 |
|  | Rh negative | None |  |  |
| All platelet transfusions |  | None |  |  |
| Platelet transfusions based on platelet count |  |  |  |  |
|  | ≤10k/μL | 2020-05-25 to 2020-05-31 | 9.84 | -5.93 — 5.41 |
|  |  | 2020-09-07 to 2020-09-13 | 10.7 | -5.93 — 5.41 |
|  |  | 2020-09-14 to 2020-09-20 | 9.73 | -5.93 — 5.41 |
|  |  | 2020-10-12 to<br>2020-10-18 | 13.5 | -5.93 — 5.41 |
|  |  | 2021-06-21 to<br>2021-06-27 | 7.11 | -5.93 — 5.41 |
|  |  | 2021-08-30 to<br>2021-09-05 | 13.7 | -5.93 — 5.41 |
|  |  | 2022-01-17 to<br>2022-01-23 | 8.58 | -5.93 — 5.41 |
|  |  | 2022-01-31 to<br>2022-02-06 | 12.4 | -5.93 — 5.41 |
|  |  | 2022-02-07 to<br>2022-02-13 | 6.55 | -5.93 — 5.41 |
|  |  | 2022-05-02 to<br>2022-05-08 | 8.63 | -5.93 — 5.41 |
|  |  | 2022-08-08 to<br>2022-08-14 | 5.66 | -5.93 — 5.41 |
|  | >10 to ≤50k/μL | None |  |  |
|  | >50 to ≤150k/μL | 2021-10-11 to<br>2021-10-17 | 13.7 | -12.1 — 11.9 |
|  |  | 2022-07-11 to<br>2022-07-17 | 14.6 | -12.1 — 11.9 |

RBC: red blood cell; *R*: remainder value; *Rlim*: remainder limits

**Figure 1.**
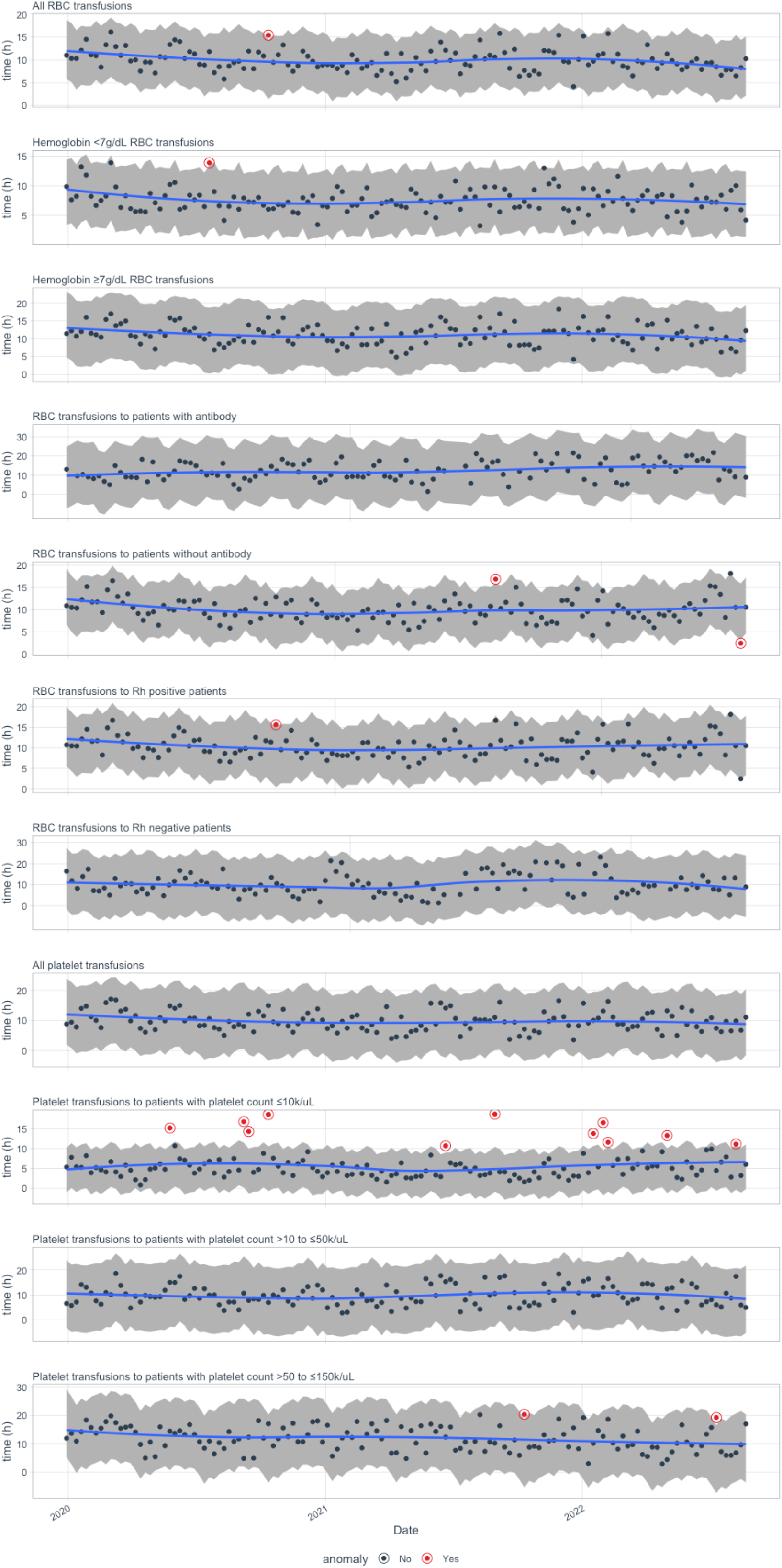
Visual representation of outlier weeks of median time to initiate a red blood cell or a platelet transfusion following a haemoglobin, or a platelet count result and based on the haemoglobin level, red blood cell minor antibody presence, Rh status, and platelet count of the patient.

Investigation of the subgroup data showed that RBC transfusions based on haemoglobin values showed only one outlier week (2020-07-20 to 2020-07-26, *R*: 6.0 [*Rlim*: -5.46 — 5.62]) for a haemoglobin value less than 7 g/dL and none for haemoglobin values ≥7 g/dL. Patients with RBC minor antibodies who received RBC transfusion did not have outliers but patients without RBC minor antibodies had two outlier weeks (2021-08-09 to 2021-08-15, *R*: 7.03; 2022-07-11 to 2022-07-16, *R*: -7.31 [*Rlim*: 6.47 — 6.13]). A single week was noted to be an outlier (2020-10-12 to 2021-08-15, *R*: 7.84 [*Rlim*: -7.38 — 7.07]) for RBC transfusions for Rh positive patients whereas no outlier weeks were noted for Rh negative patients’ RBC transfusions.

Assessment of the subgroup platelet transfusions based on pre-transfusion platelet count identified eleven weeks with outliers (2020-05-25 to 2020-05-31, *R*: 9.84; 2020-09-07 to 2020-09-13, *R*: 10.7; 2020-09-14 to 2020-09-20, *R*: 9.73; 2020-10-12 to 2020-10-18, *R*: 13.5; 2021-06-21 to 2021-06-27, *R*: 7.11; 2021-08-30 to 2021-09-05, *R*: 13.7; 2022-01-17 to 2022-01-23, *R*: 8.58; 2022-01-31 to 2022-02-06, *R*:12.4; 2022-02-07 to 2022-02-13, *R*: 6.55; 2022-05-02 to 2022-05-08, *R*: 8.63; 2022-08-08 to 2022-08-14, *R*: 5.66 [*Rlim*: -5.93 — 5.41]) for platelet count ≤10k/μL. No outlier weeks were found for platelet transfusions for patients with >10 k/μL to ≤50k/μL platelet counts, however, two weeks were identified as outliers (2021-10-11 to 2021-10-17, *R*: 13.7; 2022-07-11 to 2022-07-17, *R*: 14.6 [*Rlim*: -12.1 — 11.9]) for platelet transfusions for patients with a platelet count >50 k/μL and ≤150 k/μL.

## Discussion

Transfusion medicine service’s contingency strategies to mitigate blood shortages depends on the degree of the blood supplier’s inventory (*e*.*g*., mild: <25% lower, moderate: 25% to 50% lower, severe: 50% to 75% lower) and duration of the shortage (*e*.*g*., short term: <2 weeks, long-term: >2 weeks) and includes action items such as lowering transfusion thresholds, restricting orders, preventing wastage, splitting units, recommending pharmacological alternatives, recommending cancelation of non-essential high blood use surgeries, placing upper-limits on blood use for procedures, limiting number of available units at the remote storage refrigerators, lowering the minimum platelet yield required for the hospital to bring a unit into inventory, re-evaluating maintenance RBC exchange for sickle cell disease patients, and as a last resort extending shelf-life of blood products (6-8).

Modern machine learning (ML) tools are proposed to help manage operations by translating big data into actionable information (9). Many organizations identified successful use cases for ML tools during the throes of COVID-19 pandemic management (10). Within haematology and transfusion medicine, trend analyses and outlier detections are ML tools which could be utilized to evaluate transfusion delays. Using our proposed metric ‘weekly median time to initiate a transfusion based on an informative laboratory test result’, we have observed trends and identified numerous outliers based on: haemoglobin level, RBC minor antibody presence, Rh status, and platelet count of the patient. Although the number of outlier events were small (one for all RBC transfusions and none for all platelet transfusions), the subgroup analyses for haemoglobin <7g/dL RBC transfusions and platelet count ≤10k/μL platelet transfusions showed three and seven outlier weeks, respectively.

Delays causing major morbidity in provision of blood were noted in seven patients in the 2021 SHOT report. Five cases were included in the avoidable delays category whereas 14 cases noted in the unavoidable delays category (3). Our transfusion medicine service at Children’s National Hospital was not notified about any severe adverse events due to provision or unavailability of blood for the dates noted as outliers. We propose that the hospital transfusion medicine committees to consider time to initiate a blood transfusion from a laboratory test result which is affecting transfusion decision making (*e*.*g*., haemoglobin level or platelet count) as a metric to evaluate trends and outliers in patient’s access to blood products. This information could then be used to conduct investigations for possible missed patient adverse events and be used to address the root cause of the delays or fluctuations in transfusion timing. Furthermore, we propose conducting subgroup assessments (*e*.*g*., haemoglobin <7g/dL and platelet count ≤10k/μL transfusions), as we have observed broad analyses might miss significant events for patient subgroups.

## Data Availability

All data produced in the present study are available in de-identified fashion upon reasonable request to the authors.

